# The profile and distribution of skin disorders in ambulatory community-dwelling adult patients with Schizophrenia: a study from North India

**DOI:** 10.1101/2022.09.07.22279578

**Authors:** Natarajan Varadharajan, Tarun Narang, Seema Rani, Hitaishi Mehta, Sanjana Kathiravan, Shubh Mohan Singh

## Abstract

**Introduction:** Schizophrenia is a severe mental disorder. There is ample evidence to suggest that there are various multi-systemic co-morbidities in the patients with schizophrenia. However, data for dermatological comorbidities in these patients is scarce. This is relevant because of the common embryological basis of the central nervous system and skin, and also the disabling nature of schizophrenia itself.

**Objective:** To analyze the profile and distribution of cutaneous conditions in patients with schizophrenia.

**Methods:** Consecutive adult patients with schizophrenia attending the follow-up service of the outpatient clinic of the department of psychiatry of a tertiary hospital in North India were evaluated for the presence of a skin disorder by a dermatologist.

**Results:** Dermatological findings were seen in 71 patients (69.60%), 39 patients (38.23%) had multiple skin conditions. As a group, infections were the most prevalent diagnosis seen in 18 patients (17.64%) and fungal infections were commonly observed (n=14, 13.72%). Other common dermatoses were nevi, age related cutaneous changes, dyschromias and drug-induced acne. Among medical disorders, blood pressure was found to be elevated in 28 patients (27.45%) while sixty-four patients (69.74%) were either overweight or obese.

**Conclusion:** A high prevalence of dermatological diseases is seen in patients with schizophrenia. Both caregivers and psychiatrists managing these patients should familiarize themselves with these conditions and dermatology consultation should be sought for the prompt diagnosis and management.

## Introduction

Schizophrenia is a severe and chronic mental disorder whose exact etiology remains unknown. However various strands of evidence point to the fact that schizophrenia is a brain disorder of neurodevelopmental origin ^1^.

Embryologically, the skin shares its origin with the central nervous system (CNS) and thus it stands to reason that neurodevelopmental pathologies may be associated with skin disorders ^2^. This is reflected in phenomena such as the faulty growth of skin fibroblasts sourced from patients with schizophrenia in culture media ^3^. Secondly, immunological mechanisms that influence both the CNS and skin as in schizophrenia may contribute to skin pathology in schizophrenia ^4^. Thirdly, patients with schizophrenia may have lower levels of self-care and impairment in quality of life which may lead to poor hygiene and subsequent skin disorders such as infections. The symptoms of schizophrenia may themselves lead to behaviours such as repetitive self-harming behaviours or hand washing which may manifest as skin disorders. Finally many of the skin disorders in schizophrenia may be iatrogenic. This is commonly observed in the dermatological consequences of the use of drugs such as chlorpromazine and olanzapine ^5,6^. The interface between schizophrenia in particular and skin disorders is important as both are chronic disorders. Patients with schizophrenia have higher all cause mortality and skin disorders may contribute to the same. Skin disorders may lead to greater disability and impairment in quality of life.

Patients with schizophrenia generally have a higher prevalence of non-communicable diseases and risk factors for the same ^7^. In a study on in-patients with psychiatric disorders, 69% were found to have some or the other skin disorder ^8^. Two Indian studies also reported similar results ^9,10^. A study from Egypt in psychiatric patients also showed a high prevalence of skin disorders ^11^. These studies were conducted in patients with all kinds of psychiatric disorders and not specifically schizophrenia. Studies conducted on patients with schizophrenia are few. A Taiwanese study reported that the prevalence of at least one skin disorder in 337 patients with schizophrenia was about 50% ^12^.

Thus it can be seen that the database in this area is scarce and this is especially so in the case of community-living patients with schizophrenia. Data in this area may underline the need for more integrated services for patients with schizophrenia.

### Aim and objectives

The aim of this study was to analyze the profile and distribution of cutaneous conditions with schizophrenia.

## Materials and methods

The study protocol was approved by the Institute ethics committee. Adult patients of either gender with schizophrenia visiting the outpatient clinic of the department of psychiatry of a tertiary hospital in North India were approached for entry into the study. They had to be clinically stable, community living and on stable doses of antipsychotic medications (at least 3 months preceding the evaluation for the study). The sociodemographic, anthropometric details, diastolic and systolic blood pressure ^13^, clinical global impression ^14^ and clinical profiles of patients who consented were recorded. Hypertension was defined as per norms (systolic ≥ 140mm Hg and/or diastolic ≥ 90mm Hg) ^15^. They were then examined by a dermatologist in the same hospital to diagnose the presence and type or absence of a skin disorder. Any dermatological diagnosis was noted. Dermatological diagnoses were collapsed into categories for the purposes of analysis. Appropriate statistical tests were carried out.

## Results

102 participants (66 males and 36 females; 64.7% and 35.3% respectively) were evaluated. The sociodemographic and clinical profiles are presented in Table 1. The most common antipsychotic prescribed was olanzapine in both males and females (28 and 16 patients respectively) followed by risperidone (18 and 7 patients respectively). Thus 67.64% of the study population were on stable doses of these two antipsychotics. 9 male patients and 4 female patients were prescribed typical antipsychotics (12.74%). 6 patients were aware of being hypertensive and were on treatment for the same.

**Table 1.** Sociodemographic and clinical profile of study population.

|  | Male (n=66) | Female (n=36) | p |
| --- | --- | --- | --- |
| Mean age in years (SD) | 39.30 (12.12) | 39.63 (10.60) | 0.88* |
| Mean years of education (SD) | 10.34 (3.79) | 10.08 (5.22) | 0.76* |
| Married | 35 (53.03%) | 29 (80.55) | 0.07** |
| Rural background | 42 (63.63) | 18 (50) | 0.18** |
| Mean duration of schizophrenia in months (SD) | 142.53 (108.79) | 119.02 (92.67) | 0.27* |
| Mean BMI (SD) | 25.10 (4.57) | 26.02 (5.34) | 0.36* |
| Mean waist circumference in cm (SD) | 93.87 (10.76) | 94.67 (15.10) | - |
| Hypertension | 19 (28.78) | 9 (25) | 0.68* |

P significant if <0.05, \*= Independent-samples T test, \*\*=Chi-square test, n= number, SD= standard deviation

The prevalence of obesity (body mass index≥25) was 54.90% (n=56, male=32, female=24). The prevalence of obesity in patients prescribed olanzapine was 56.81%, 48% in risperidone, 85.71% in clozapine and 83.33% in patients on injectables. However the prevalence of obesity was not statistically different in patients prescribed different antipsychotics (p=0.23).

A majority of patients were considered normal or borderline mentally ill at time of assessment (51 males and 29 females) and very much improved or much improved due to drug treatment (60 males and 32 females) as per the clinical global impression ^14^.

### Dermatological disorders

The prevalence of any dermatological finding was 69.60% (n=72, male=47, female=25, p=0.85), 45 patients (63.38% of those with dermatological diagnosis) had more than one dermatological diagnosis. The total number of diagnoses made were 136 and 71 different diagnoses were noted. We analysed all the diagnoses made. Table 2 presents the most common dermatological diagnoses in the participants (>1 mention). Table 3 mentions the most common categories of diagnoses.

**Table 2.** Most common diagnoses (>1 mention)

| Diagnosis | n |
| --- | --- |
| Melasma | 9 |
| Acne Vulgaris | 9 |
| Cherry angioma | 8 |
| Melanocytic nevi | 8 |
| Onychomycosis | 6 |
| Freckles | 6 |
| Acanthosis nigricans | 4 |
| Rosacea | 3 |
| Tinea cruris | 3 |
| Polymorphous light eruption | 3 |
| Beau's lines | 3 |
| Folliculitis | 3 |
| Melanonychia | 3 |
| Post varicella scarring | 3 |
| Xerosis | 3 |
| Androgenic alopecia | 2 |
| Dermatitis passivita | 2 |
| Tinea corporis | 2 |
| Post Inflammatory hyperpigmentation | 2 |
| Lichen simplex | 2 |
| Seborrheic keratosis | 2 |
| Psoriasis | 2 |

n= number

**Table 3.**
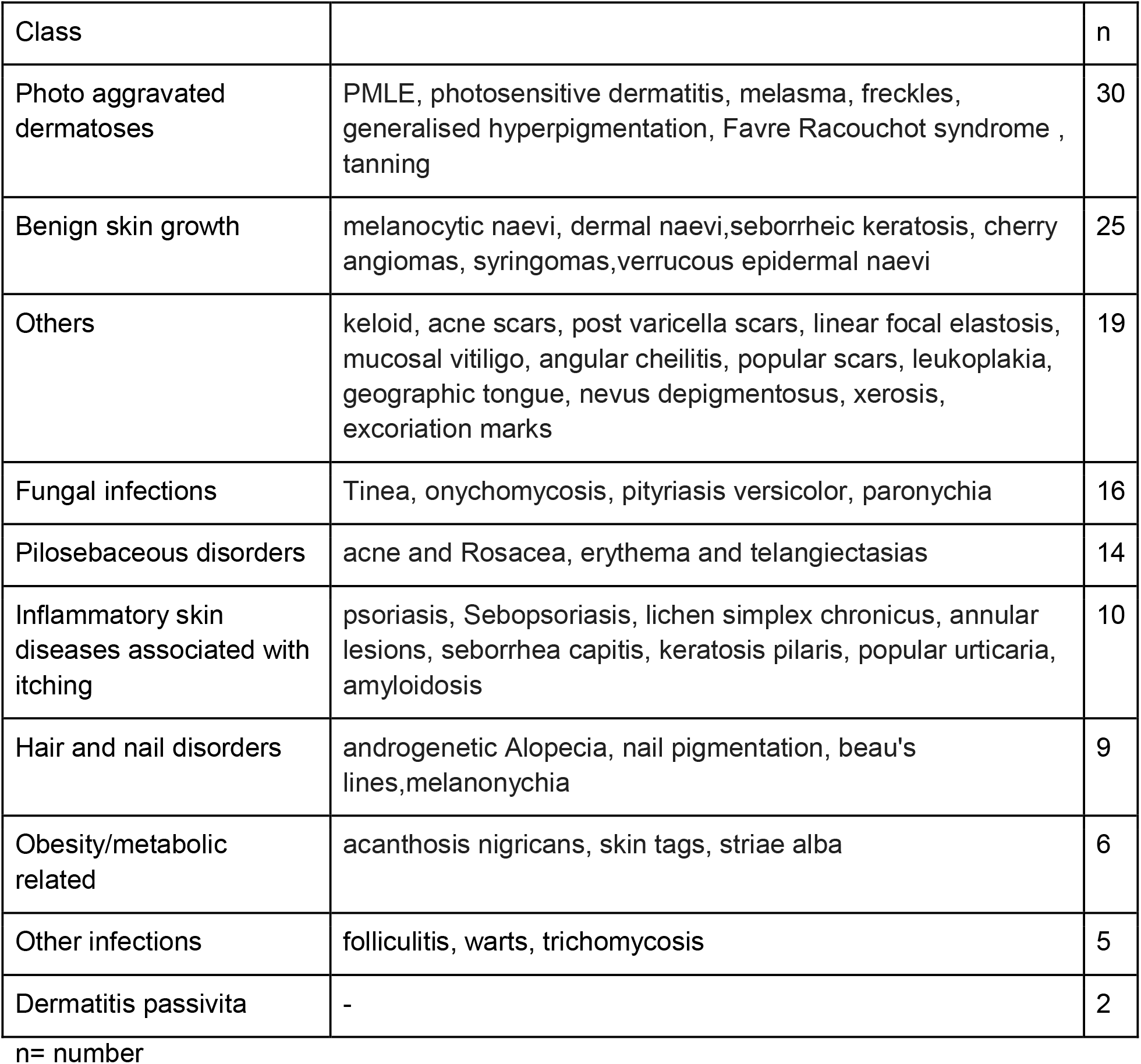
Most common classes of dermatoses.

| Class |  | n |
| --- | --- | --- |
| Photo aggravated dermatoses | PMLE, photosensitive dermatitis, melasma, freckles, generalised hyperpigmentation, Favre Racouchot syndrome , tanning | 30 |
| Benign skin growth | melanocytic naevi, dermal naevi, seborrheic keratosis, cherry angiomas, syringomas, verrucous epidermal naevi | 25 |
| Others | keloid, acne scars, post varicella scars, linear focal elastosis, mucosal vitiligo, angular cheilitis, popular scars, leukoplakia, geographic tongue, nevus depigmentosus, xerosis, excoriation marks | 19 |
| Fungal infections | Tinea, onychomycosis, pityriasis versicolor, paronychia | 16 |
| Pilosebaceous disorders | acne and Rosacea, erythema and telangiectasias | 14 |
| Inflammatory skin diseases associated with itching | psoriasis, Sebopsoriasis, lichen simplex chronicus, annular lesions, seborrhea capitis, keratosis pilaris, popular urticaria, amyloidosis | 10 |
| Hair and nail disorders | androgenetic Alopecia, nail pigmentation, beau's lines, melanonychia | 9 |
| Obesity/metabolic related | acanthosis nigricans, skin tags, striae alba | 6 |
| Other infections | folliculitis, warts, trichomycosis | 5 |
| Dermatitis passivita | - | 2 |

n= number

There was no statistical difference between those with and without dermatoses with regards to gender (p=0.81), obesity (p=0.83), clinical state (p=0.32) or improvement due to psychotropic drugs (p=0.51).

## Discussion

Skin disorders are known to be associated with psychiatric disorders including schizophrenia (15,16). Studies dealing with skin disorders in schizophrenia include inquiries into specific disorders such as psoriasis or into the general prevalence of any skin disorder in schizophrenia (15). The latter type of studies are scarce (12). These studies are relevant from a clinical perspective because schizophrenia is known to be associated with a high degree of comorbidity with various non-communicable and communicable diseases. Thus this study was carried out to add to the literature base in this area. The results of this study must be discussed with reference to the participants being health-care seeking patients in a tertiary hospital, the absence of a control group, the possibility of a selection bias in that patients with skin complaints may have consented with greater alacrity and a modest sample size. However, most participants did not volunteer any skin complaints and the findings are a product of careful dermatological examination.

The socio-demographic and clinical profile of the study group was fairly typical of the population of treatment-seeking, community-living and stable patients with schizophrenia visiting the outpatient clinic of the department of psychiatry. The prevalence of hypertension, obesity and abdominal obesity is also similar to that reported earlier from this centre ^16^.

Our study indicates that there is a high prevalence of skin disorders in patients with schizophrenia. This is similar though slightly less than that found in a study conducted in Taiwan (12). The prevalence of skin disorders in this population is also higher than that has been observed in the general population in rural India ^17^ but less than that found in clinic based studies of patient populations such as those with diabetes ^18^. As has been pointed out earlier, there is a paucity of systematic studies with regards to skin disorders in schizophrenia and thus comparisons are difficult for this reason ^19^.

With regards to our findings, the following observations can be made. The most common group of disorders was that of photo aggravated dermatoses (29.41%). While photosensitivity has been recognised as a side-effect of typical antipsychotics, it is increasingly being recognised as being frequent with the use of newer atypical antipsychotics as well ^20^. Interestingly enough, the study from Taiwan did not report photosensitivity reactions in it’s sample ^12^. Photosensitivity reactions can have various manifestations and severities. Our study shows that in an Indian population with schizophrenia and on antipsychotic medications, photosensitivity reactions are common. These disorders need to be identified and treated early as pigmentary disorders on the face are associated with significant impairment of quality of life and require long term management.

Benign skin growths are non-cancerous and are frequently observed in clinical and non-clinical populations. These were the second most prevalent group of skin disorders in our study population. These are observed in all cultures and across all age groups. The main point of importance is that they require to be differentiated from malignant lesions ^21^. Other disorders such as keloids, acne scars, post varicella scars, linear focal elastosis, mucosal vitiligo, angular cheilitis, popular scars, leukoplakia, geographic tongue, nevus depigmentosus, xerosis, excoriation marks were also observed in our study population. Also patients with inflammatory skin diseases, and hair and nail disorders were commonly observed in the study group. These are likely to be incidental and unrelated to the effects of schizophrenia or its treatment ^12^.

Infections (fungal and non-fungal) were common in our patient population (24.50%). The prevalence of infections in this population can have various interacting etiologies. These can range from immune abnormalities, metabolic syndrome and altered glycemic control, altered skin microbiome and general lack of hygiene due to the effects of the illness ^22^. Also disorders like tinea or onychomycosis need to be treated,as they are associated with significant morbidity and transmission to family members also. The treatment is systemic antifungals is expensive and if the condition is not treated early and adequately it may become more severe, which requires a longer duration of treatment.

The prevalence of obesity associated skin disorders was 10.71% in the population found to be obese. As has been pointed out, obesity is a common comorbidity in schizophrenia and has multi-factorial origin ^16^. Our study shows the importance of assessment of this group of dermatoses in patients with schizophrenia who are also obese.

The association of pilosebaceous disorders especially acne and antipsychotic medications is well known ^12^.This relationship is complex and multifactorial. These disorders are especially troublesome in schizophrenia given their propensity to cause disfigurement ^23^.

We detected two patients with dermatitis passivita ^24^. This perhaps reflects the overall clinical status of the study group in that schizophrenia in most patients was well controlled and they were on stable doses of medication. It is possible that the dermatological manifestations in patients with more severe or poorly controlled schizophrenia may be different and possibly more severe.

## Conclusion

Our study emphasizes the importance for psychiatrists to be aware of the high comorbidity between schizophrenia and skin disorders.

## Data Availability

All data produced in the present study are available upon reasonable request to the authors

## Acknowledgments

None

